# Machine learning-based prediction of response to PARP inhibition across cancer types

**DOI:** 10.1101/19007757

**Authors:** Katherine E. Hill, Ahmed Rattani, Christopher E. Lietz, Cassandra Garbutt, Edwin Choy, Gregory M. Cote, Aedin Culhane, Andrew D. Kelly, Benjamin Haibe-Kains, Dimitrios Spentzos

## Abstract

PARP inhibitors (PARPi) are FDA approved for the treatment of BRCA1/2 deficient breast and ovarian cancer, but a growing body of pre-clinical evidence suggests the drug class holds therapeutic potential in other cancer types, independent of BRCA1/2 status. Large-scale pharmacogenomic datasets offer the opportunity to develop predictors of response to PARPi’s in many cancer types, expanding their potential clinical applicability. Response to the PARPi olaparib was used to identify a multi-gene PARPi response signature in a large *in vitro* dataset including multiple cancer types, such as breast, ovarian, pancreatic, lung cancer, osteosarcoma and Ewing sarcoma, using machine learning approaches. The signature was validated on multiple independent *in vitro* datasets, also testing for response to another PARPi, rucaparib, as well as two clinical datasets using the cisplatin response as a surrogate for PARPi response. Finally, integrative pharmacogenomic analysis was performed to identify drugs which may be effective in PARPi resistant tumors. A PARPi response signature was defined as the 50 most differentially transcribed genes between PARPi resistant and sensitive cell lines from several different cancer types. Cross validated predictors generated with LASSO logistic regression using the PARPi signature genes accurately predicted PARPi response in a training set of olaparib treated cell lines (80-89%), an independent olaparib treated *in vitro* dataset (66-77%), and an independent rucaparib treated *in vitro* dataset (80-87%). The PARPi signature also significantly predicted *in vitro* breast cancer response to olaparib in another separate experimental dataset. The signature also predicted clinical response to cisplatin and survival in human ovarian cancer and osteosarcoma datasets. Robust transcriptional differences between PARPi sensitive and resistant tumors accurately predict PARPi response *in vitro* and cisplatin response *in vivo* for multiple tumor types with or without known BRCA1/2 deficiency. These signatures may prove useful for predicting response in patients treated with PARP inhibitors.

## Introduction

Genome fidelity is essential for the survival of cells and organisms. In addition to the intrinsic proofreading exonuclease activities of the DNA polymerase and replication complexes, eukaryotic cells have evolved elaborate error correction pathways able to repair both single and double strand damages [1]. Germline or somatic mutations in the genes involved in DNA repair pathways, such as BRCA1 and BRCA2, are frequently observed in cancer cells [2]. Cells with defects in the BRCA1 or BRCA2 pathways, which impair double strand break (DSB) repair, rely on single strand repair pathways, such as base excision repair pathway (BER), to correct frequently occurring DNA damage.

The poly ADP ribose polymerase (PARP) protein family, which includes PARP1 and PARP2, are a vital component of BER pathway [3],[4]. In addition to its role in BER repair, PARP1 also participates in DSB repair via activation of ATM, a protein necessary for homologous recombination (HR) repair [5]. PARP1 also recognizes stalled replication forks and recruits protein to begin HR repair [6]. These functions make the PARP family an appealing target in cancer cells, especially those exhibiting mutations in double strand repair pathway [7].

Cancer cells with BRCA and other homologous recombination pathway mutations are exquisitely sensitive to PARP inhibitors [8],[9], and patients with ovarian cancer [10], breast cancer [11], [12], pancreatic cancer [13], and prostate cancer [14] with germline BRCA mutations have improved progression-free survival on these medications [15]. Though germline and somatic BRCA mutation patients derive the most benefit from these drugs, in clinical trials, the survival advantage of PARP inhibitors also extends to a subset of wild type BRCA tumors [16], which are often HR deficient by other means such as PALB2 mutation or BRCA promoter methylation. Moreover, emerging cell line and early-stage clinical trial data indicate that PARP inhibition might offer clinical benefits in cancers not typically associated with BRCA mutations, for example, in prostate, lung, Ewing sarcoma, osteosarcoma, and BRCA wild type breast cancer. Thus, there is a need to develop tumor subtype agnostic biomarkers which reliably predict clinical response to PARP inhibitors.

We, therefore, set out to develop a PARP response signature using gene expression data from a very large collection of cell lines treated with the PARP inhibitor olaparib, the first such drug that entered clinical development [17]. We developed a predictor using machine learning algorithms and validated it on independent groups of cell lines treated with both olaparib and rucaparib. The model’s predictive value was further independently validated in three additional data sets, two of which were clinical, using cisplatin response as a surrogate. Further biologic and therapeutic implications of our findings are discussed.

## Results

### Highly predictive gene signature for PARP inhibitor response

We asked if the in-vitro sensitivity to olaparib and rucaparib (previously called AG-014699) of tumor cell lines from a wide range of tissue subtypes previously reported in a large pharmacogenomic study [18] is also associated with transcription patterns, which could then be used as markers for PARP inhibitor response prediction across many different tissue subtypes. We also explored if we could develop a multi-gene panel predictive of PARP responsiveness in patient tumors and cells lines without any discernible genetic hallmarks of BRCA gene or related pathway deregulation.

To minimize both technical laboratory artifact effects and also to pursue “drug class” as opposed to drug specific findings, we performed our analysis only in the large subset of cell lines, (n=143) that showed concordant sensitivity or resistance to the two drugs (Supplementary Table 1). We first identified differentially expressed genes in a subset of cell lines treated with olaparib. Due to the large number of statistically significantly differentially expressed genes (Supplementary Table 2), we selected the top 50 genes and used their expression levels to classify in-vitro response of cell lines to PARP inhibitors. For a training set, we randomly selected two thirds of the cell lines treated with olaparib, and one third was used for initial validation.

**Table 1.** Genes selected by LASSO logistic regression model for PARPi response prediction in the cell line training dataset by Garnett et al.

| Gene symbol | Gene name | Coefficient |
| --- | --- | --- |
| CALM1 | calmodulin 1 | -0.86939 |
| CALM1 | calmodulin 1 | -0.34952 |
| MMP2 | matrix metalloproteinase 2 | -1.28786 |
| NID1 | nidogen 1 | 2.96661 |
| PDGFRB | platelet-derived growth factor receptor beta | 2.73977 |
| TP53BP1 | tumor protein p53 binding protein 1 | 1.09617 |
| GALNT3 | polypeptide N-acetylgalactosaminyltransferase 3 | 1.98191 |
| PRAF2 | PRA1 domain family, member 2 | -1.36475 |
| COX11 | cytochrome c oxidase copper chaperone COX11 | 4.19121 |
| HTATIP2 | HIV-1 Tat interactive protein 2, 30kDa | 0.00954 |
| MYD88 | myeloid differentiation primary response 88 | -4.33323 |
| HTATIP2 | HIV-1 Tat interactive protein 2, 30kDa | -0.35708 |
| CALM1 | calmodulin 1 | -2.88417 |
| MAPK13 | mitogen-activated protein kinase 13 | 0.42046 |
| PXDN | peroxidasin | -0.42148 |
| SOS2 | SOS Ras/Rho guanine nucleotide exchange factor 2 | -3.04141 |
| RUFY1 | RUN and FYVE domain containing 1 | 5.16040 |
| NLRP1 | NLR Family Pyrin Domain Containing 1 | 1.02986 |
| HCFC1R1 | host cell factor C1 regulator 1 | 0.05239 |
| LARP6 | La ribonucleoprotein domain family, member 6 | 1.91950 |
| CNTNAP1 | contactin associated protein 1 | 1.82016 |
| CLSTN2 | calsyntenin 2 | 1.20249 |

**Table 2.** PARP response signature genes included in optimal logistic regression model for cisplatin response prediction in the ovarian cancer data.

| Gene symbol | Gene name | Coefficient |
| --- | --- | --- |
| MMP2 | matrix metalloproteinase 2 | 0.21011 |
| PKIG | protein kinase inhibitor gamma | 0.07603 |
| TP53BP1 | tumor protein p53 binding protein 1 | 0.11472 |
| PRAF2 | PRA1 domain family, member 2 | 0.12707 |
| HTATIP2 | HIV-1 Tat interactive protein 2, 30kDa | 0.10122 |
| HTATIP2 | HIV-1 Tat interactive protein 2, 30kDa | 0.05653 |
| CALM1 | calmodulin 1 | -0.01888 |
| MAPK13 | Mitogen-activated protein kinase 13 | -0.17067 |
| ITPK1 | inositol 1,3,4-triphosphate 5/6 kinase | -0.25138 |
| HCFC1R1 | Host cell factor C1 regulator | 0.19645 |

The model derived from the training set was then applied to cell lines treated with rucaparib for another independent testing. On 3-fold cross validation LASSO-logistic regression, prediction accuracies with various subsets of the 50 genes ranged from 80-89% for the training set, 66-77% for the olaparib-treated independent cell lines and 80-87% for the rucaparib treated independent data set. An example of a high performing logistic regression-based prediction model is shown in Table 1.

On inspection of classification accuracies, we found our prediction algorithm worked very well for certain tissue subtypes, such as ovarian, and breast cancer cells, which were classified with 100% accuracy. That said, the signature also showed 100% cross validated prediction accuracy in osteosarcoma cancer cell lines and 71% and 84% for Ewing’s and the lung cancer cell lines, respectively, observations that are interesting given the lack of any clear knowledge for genetic perturbations of the HR pathway in any significant subsets of these tumors. We noted that there is substantial heterogeneity in PARP inhibitor drug response between the different tissue types included in the Garnett dataset, with some cell types showing extreme rates of resistance or sensitivity (approaching 100%), and others showing a more “balanced” split in terms of the fraction of sensitive or resistant cell lines. Therefore, we considered the possibility that this type of “tissue bias” might mean that the gene expression differences and model predictions may reflect to a large extent differences between the different histologic cancer types as opposed to a true PARPi resistance phenotype. To answer this question, we assessed the classification accuracy excluding any cell lines from tissue types that showed above 80% resistance or sensitivity to either of the two drugs (Supplementary Table 1). We observed that the prediction accuracy in the remaining cell lines (n = 111) was similar, 80-89%, 68-77%, and 79-86% for the training, independent olaparib and the independent rucaparib treated datasets, respectively.

We then asked if a published ovarian cancer “BRCAness” signature [19], that was previously shown by our group to predict response to PARP inhibitors in ovarian tumors that did not harbor BRCA mutations, could also be applied broadly to the multiple different tissue subtypes included in the current analysis. The “BRCAness” signature, when used for hierarchical clustering [20] of the cell lines, showed a significant association with *in-vitro* response to PARP inhibitors (Fisher’s exact test p-value: < 0.05; odds ratio (OR): 2.1 - 3.9). However, a 3-fold cross-validated LASSO logistic regression prediction model showed accuracies only in the range of 56 - 72%. This suggests that this “BRCAness” signature, while associated with defects in the DNA repair pathway in tumors that do not harbor BRCA mutations, does not fully capture the biology that determines response to PARP inhibitors in different tissue subtypes, underscoring the possible new insights offered by our current analysis.

Finally, we applied the PARPi response signature on an entirely separate public dataset from a different study [21] of gene expression profiles of 9 breast cancer cell lines treated with olaparib. Hierarchical clustering using PARP inhibitor response gene lists ranging from 2 to 49 genes from our signature distinguished between the relatively resistant and sensitive cell lines in this external dataset (OR ≥ 4.0; P = 0.008 - 0.048; Figure 1A). Least angle regression (LARS) models significantly predicted each cell line’s response to olaparib. A representative model is shown in Figure 1B (8 gene model, 0% deviation from minimal error allowed, R^2^ = 0.77).

**Figure 1.**
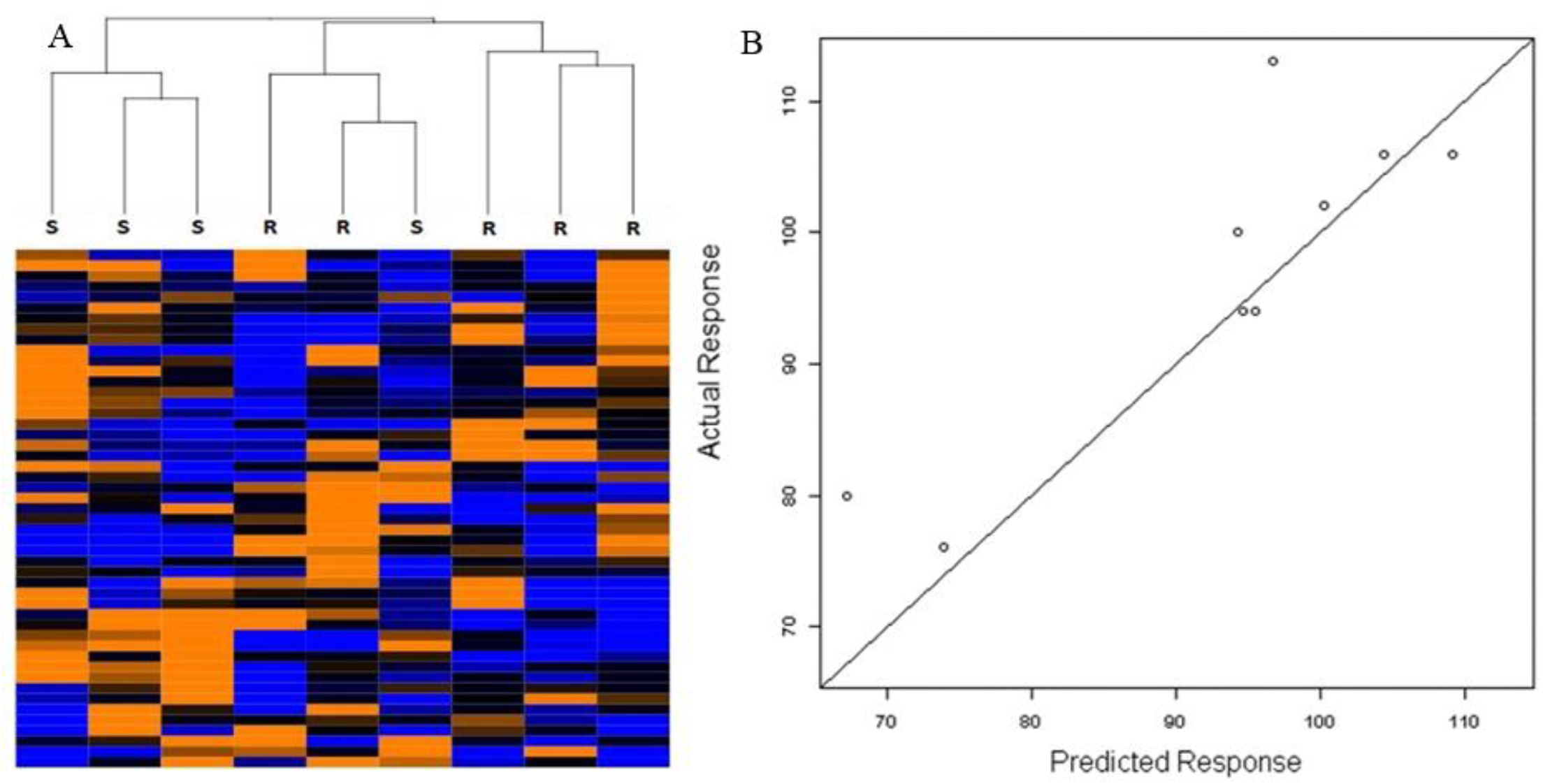
A): Hierarchical clustering and Heat map with the 50 PARP inhibitor response signature transcripts in an independent breast cancer cell line dataset separate from the GCSD dataset shows good discrimination between resistant (R) and sensitive (S) cell lines (Fisher’s p = 0.048). Separation was even more significant with smaller gene subsets. B): LARS regression response prediction for individual cell lines with an 8 gene subset of PARP inhibitor response genes in the same breast cancer cell line dataset.

### Assessment of individualized prediction accuracy via ROC analysis

Receiver Operator Characteristic (ROC) analysis is the gold standard for assessing predictive power of a given biomarker on a continuous variable scale. Thus, we performed ROC analysis in the independent validation set of 143 cell lines treated with rucaparib. The 22-gene predictor model presented above in Table 1, achieved an AUC = 0.919 (95% CI: 0.87 - 0.97, p < 0.001, Figure 2). Various models including subsets of 14 - 50 genes all performed similarly with statistically significant AUC values 0.88 - 0.90.

**Figure 2.**
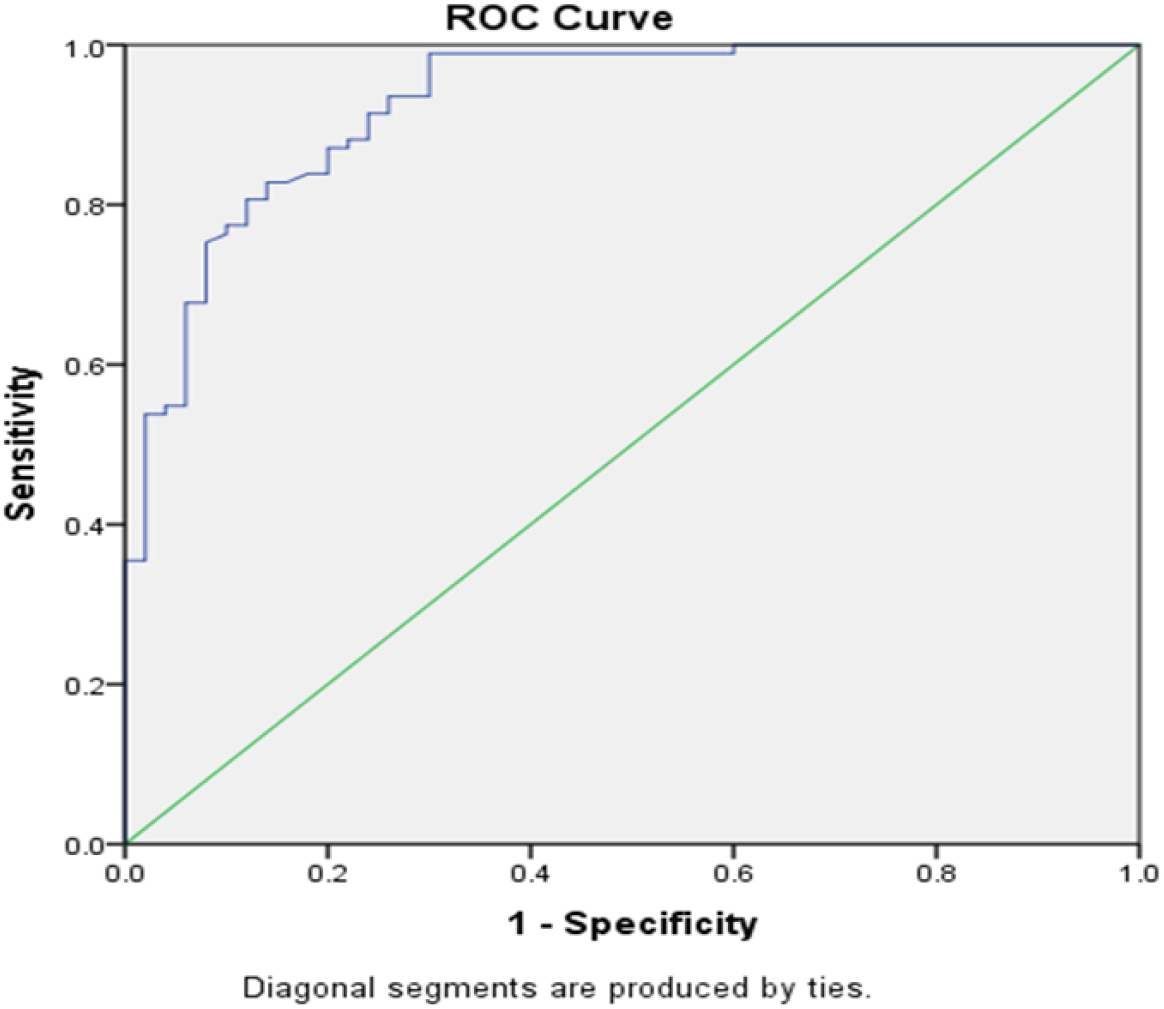
ROC assessment of the performance of the gene expression predictor in the independent rucaparib-treated cell line dataset. AUC = 0.927 (95% CI: 0.88 - 0.97, p < 0.001)

### PARP response signature is associated with clinical chemo response and recurrence in ovarian cancer

To test if the PARP response signature identified *in vitro* could be used to determine the in-vivo tumor sensitivity, we selected 456 cases of ovarian cancer treated with cisplatin from The Cancer Genome Atlas network dataset [22]. Given the lack of tumor derived molecular data from patients treated with PARP inhibitors to date, cisplatin response was analyzed as a surrogate for sensitivity to agents targeting DNA repair pathways [19], and specifically PARPi’s. A 3-fold cross-validated LASSO-logistic regression models constructed iteratively with 25 - 50 gene subsets yielded accuracies of approximately 68%, in a training set including 2/3 of the cases, while predictions in the independent sample set including the remaining 1/3 of samples ranged between 70 - 74%. Genes from the optimal cisplatin prediction model in the ovarian cancer data are seen in Table 2. In order to account for possible overfitting confounding these findings we also performed unspervised hierarchical clustering and found a significant association between PARPi response signature derived cluster groups and clinical response in the TCGA samples (Fisher’s exact test P = 0.038 - 0.076, OR = 1.46 - 1.60) (Figure 3), further indicating that the PARPi response profiles do carry biologic relevance to cisplatin response in a clinical cohort.

**Figure 3.**
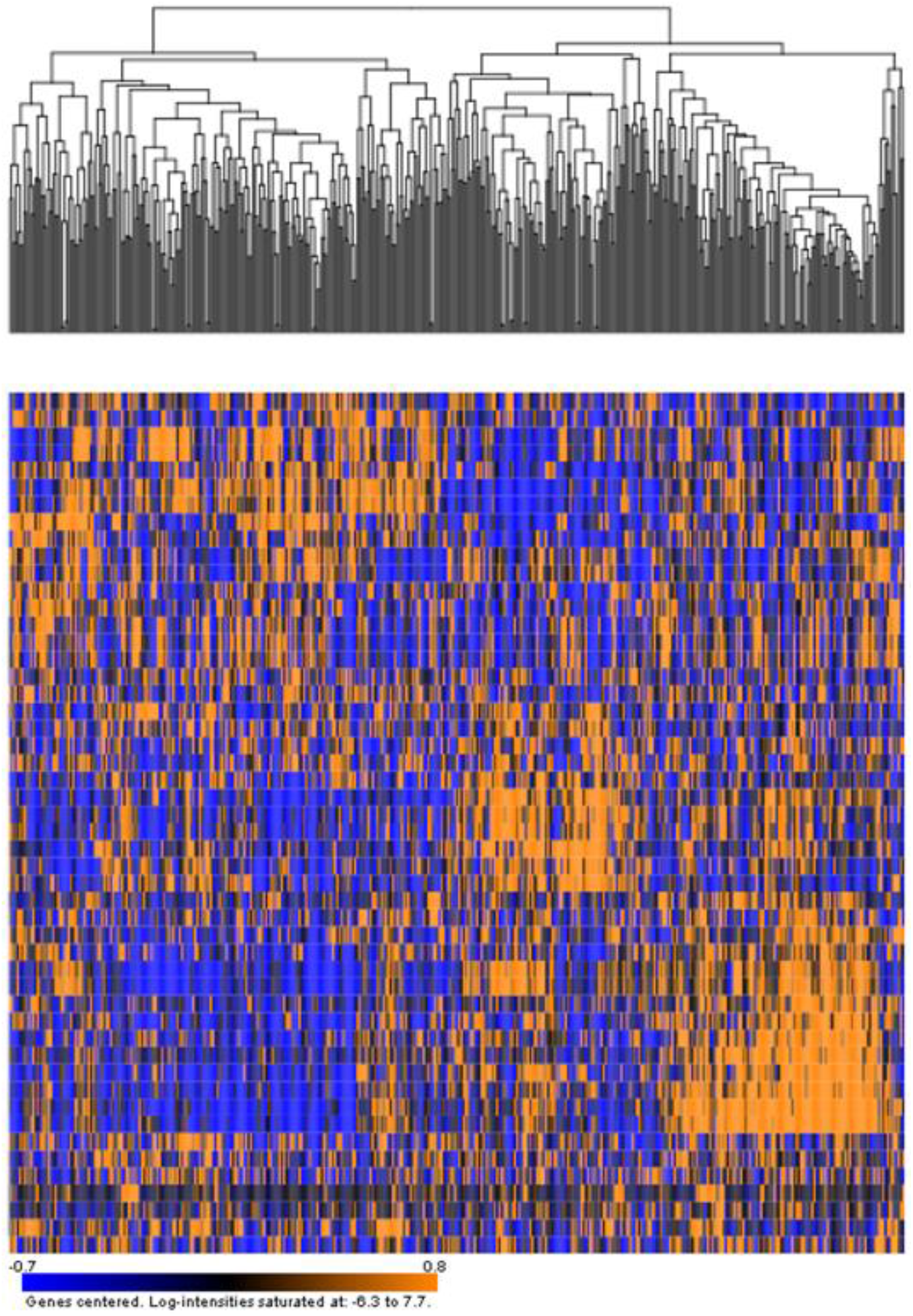
Hierarchical clustering and heatmap using the 50 gene PARP inhibitor response signature in the human TCGA ovarian cancer dataset. The two main dendrogram branches were significantly associated with cisplatin response.

We further found that the PARP inhibitor response signature not only predicted in-vivo response of the tumor to cisplatin, but was also associated with recurrence free survival in the ovarian cancer clinical data. We examined the iteratively generated cluster groups described above, based on unsupervised hierarchical clustering with several parsed subsets of the 50 differentially expressed mRNAs, and found that they also represented two groups with different recurrence free survival risk. For example, Figure 4 shows Recurrence Free Survival results based on a 40-gene derived sample clustering, (Log-rank P < 0.001; Hazard Ratio (HR): 1.75; 95% confidence interval (CI): 1.30 - 2.27; Median RFS: 551 months vs. 843 months); and a 14-gene derived clustering (Log-rank P = 0.006; HR = 1.44; 95% CI: 1.11 - 1.87; Median RFS: 552 months vs. 680 months;). Supervised models using the same gene lists also predicted groups with recurrence free survival that was very close to nominal signifiance (14 gene model, Log-rank P = 0.060; Permutation P = 0.17; HR: 1.275; 95% CI: 0.990 - 1.643; Medan RFS: 582 months vs. 646 months; Figure 4). PARP response genes with the strongest univariate association with RFS are shown in Table 3.

**Table 3.** PARP response signature genes associated with clinical outcome (RFS) in ovarian cancer.

| Gene symbol | Gene name | Hazard ratio | Parametric p-value |
| --- | --- | --- | --- |
| CALM1 | calmodulin 1 | 0.744 | 0.017 |
| CALM1 | calmodulin 1 | 0.766 | 0.018 |
| KCNJ4 | potassium inwardly-rectifying channel, subfamily J, member 4 | 1.327 | 0.022 |
| TP53BP1 | tumor protein p53 binding protein 1 | 1.315 | 0.026 |
| MAPK13 | mitogen-activated protein kinase 13 | 0.840 | 0.028 |
| ITPK1 | inositol 1,3,4-triphosphate 5/6 kinase | 0.792 | 0.046 |
| LDB2 | LIM domain binding 2 | 1.203 | 0.047 |
| CALM1 | calmodulin 1 | 0.761 | 0.052 |
| HCFC1R1 | host cell factor C1 regulator 1 | 1.170 | 0.076 |
| PKIG | protein kinase inhibitor gamma | 1.158 | 0.092 |
| CNTMAP1 | contactin associated protein 1 | 1.463 | 0.092 |

**Figure 4.**
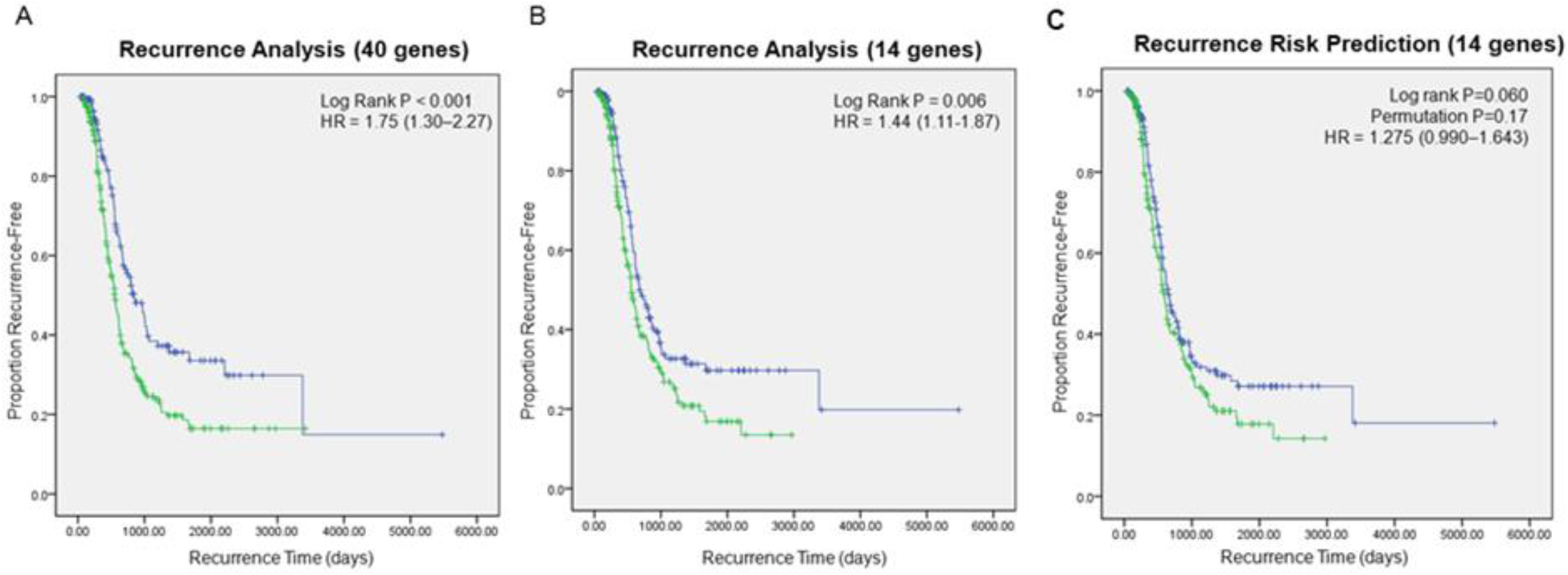
Recurrence Free Survival analysis with PARP inhibitor response genes in TCGA ovarian cancer samples. A) Kaplan-Meier RFS analysis based on unsupervised hierarchical clustering with 40 genes B) Kaplan-Meier RFS analysis based on unsupervised hierarchical clustering with 14 genes. C) Supervised prediction model for RFS with 14 genes.

### PARP response signature is associated with clinical chemoresponse and recurrence in osteosarcoma

Osteosarcoma is one of the most recalcitrant cancers with five-year survival rates for recurrent and metastatic cases below 30% and inadequate response to chemotherapy beyond first line. Because of the unexpected response to PARP inhibitors also observed in a subset of osteosarcoma cell lines (Supplementary Table 3), we tested if the PARP inhibitor response signature we identified using the gene expression of cell lines exposed to olaparib could be useful in the clinical setting. For this, we used gene expression data that we previously published from a human cohort of 33 primary osteosarcoma samples [23]. As we did in the ovarian cancer analysis, and given that cisplatin is part of the standard first line treatment in osteosarcoma, we also used cisplatin response as a surrogate for PARP inhibitor response.

We first mapped our signature probe sets (which was generated on the Affymetrix platform) to the Illumina DASL platform which was used in the previous human osteosarcoma gene expression study and found that 46 of the 50 genes could be mapped.

The low sample size of the osteosarcoma clinical cohort and differences in microarray platforms, presented special challenges for this analysis. Despite these limitations we did find evidence that the PARP inhibitor signature was associated with chemoresponse assessed by cisplatin induced tumor percent necrosis in the osteosarcoma cohort. Among the signature genes, in univariate logistic regression analysis COX11, EDNRA, and LDB2 were significantly associated with percent tumor necrosis (P = 0.033, 0.080, 0.067, ORs: 1.89, 2.41, 2.11 respectively, > 90% vs < 90%), while COX11, RUFY1, and LDB2 were significantly associated with tumor percent necrosis (Pearson: 0.338, 0.356, −0.405; P = 0.055, 0.042, 0.02, respectively). LASSO logistic regression prediction models (LOO cross-validated) using from 2 to 50 PARP response genes predicted clinical response to cisplatin with accuracies ranging 61-73%. Further, 7 mRNAs from the PARP inhibitor response signature were found to be associated with RFS by standard univariate Cox proportional hazards models with a significant or trending p value (P < 0.15; Table 4). Finally, unsupervised hierarchical clustering based iteratively on 10-30 PARP response signature gene subsets demonstrated differences in recurrence free survival (for example, 30 genes: median RFS: 14 months versus 151 months, HR = 6.34, 95% CI: 2.00-20.53, log-rank P < 0.001, Figure 5A; 10 genes: median RFS: 34 months versus not-yet-reached, HR = 2.09, 95% CI: 0.76-5.75, log-rank P = 0.003, Figure 5B). Given the substantial technical differences between the datasets and the small sample size of the osteosarcoma cohort, we find these results supportive of the general hypothesis that the PARP response signature may be clinically applicable in osteosarcoma too.

**Table 4.** PARP response signature genes associated with clinical outcome (RFS) in osteosarcoma.

| Gene symbol | Gene name | Hazard ratio | Parametric p-value | Standard deviation of log intensities |
| --- | --- | --- | --- | --- |
| PDGFRB | platelet-derived growth factor receptor beta | 1.722 | 0.044 | 1.500 |
| LDB2 | LIM domain binding 2 | 1.413 | 0.049 | 1.195 |
| CALM1 | calmodulin 1 | 1.813 | 0.058 | 0.995 |
| CNTNAP1 | contactin associated protein 1 | 1.473 | 0.074 | 1.340 |
| MRC2 | mannose receptor C type 2 | 1.373 | 0.106 | 1.669 |
| ITPK1 | inositol 1,3,4-triphosphate 5/6 kinase | 0.701 | 0.111 | 1.452 |
| DENND5A | DENN domain containing 5A | 1.472 | 0.116 | 0.827 |

**Figure 5.**
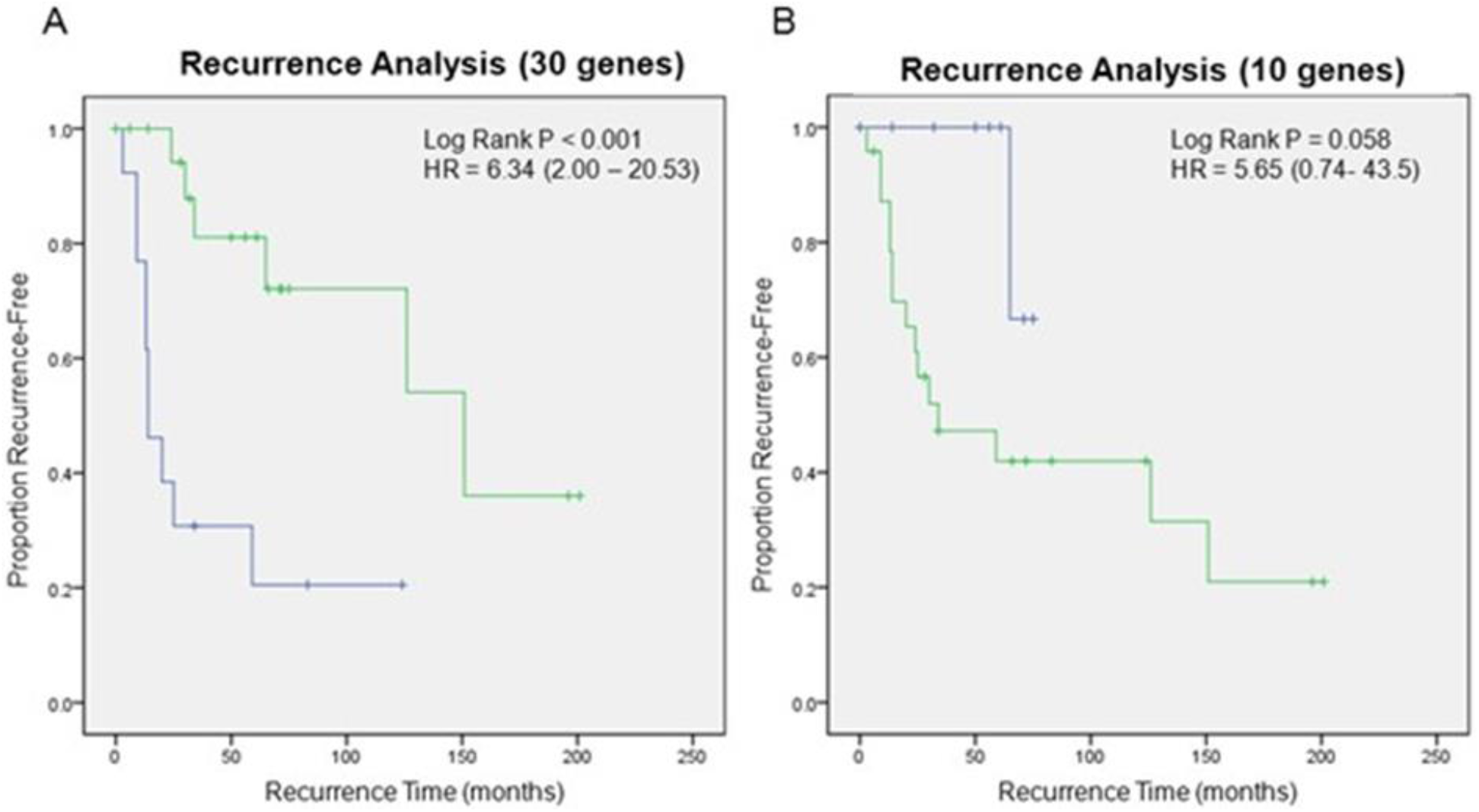
Recurrence Free Survival analysis with PARP inhibitor response genes in an osteosarcoma cohort. A) Kaplan-Meier RFS analysis based on unsupervised hierarchical clustering with 30 genes B) Kaplan-Meier recurrence analysis based on unsupervised hierarchical clustering with 10 genes.

### Gene Set Enrichment analysis demonstrates critical biologic processes and pathways differentially regulated in the PARP resistant versus sensitive phenotype

The substantial global differences in transcription between PARPi sensitive and resistant cell lines prompted us to investigate if specific functional pathways are implicated in PARPi response. We performed Gene Set Enrichment Analysis and identified many pathways differentially regulated by virtue of coordinated/aggregated differences of the pathway gene sets between cell lines sensitive or resistant to olaparib and rucaparib (LS and KS p < 0.05) (Table 5 and Supplementary Table 4). This analysis was performed using the global expression data filtered only by 20% for low variance genes (analysis with 10% filter and no filter produced very similar results). Predefined signatures of oncogenic pathway activation associated with PARPi response included pathways which may be therapeutically targetable, such as MEK and mTOR, among others. Expression of genes located on the largest non-coding cluster in the human genome, 14q32, were found to be significantly different between PARPi resistant and sensitive cell lines. Transcription and epigenetic modification of this locus has been shown to predict outcome in various cancer type by our group and others [23],[24],[25]. This finding is supported by differential expression of the gene target set of miR-299, a microRNA that is also located on 14q32

**Table 5.** Gene Set Analysis. Gene sets with LS and KS permutation p < 0.05 when analyzed in both the olaparib and rucaparib datasets were considered enriched. The sign of the Efron – Tibshirani maxmean statistic indicates if the geneset is over (+) or under (-) expressed in PARPi sensitive cell lines. In the large majority of gene sets, the direction of differential regulation was concordant between the olaparib and rucaparib treated cell lines, with only a few exceptions, in which case a sign is not listed.

| Cytoband |  | Pathways |  | microRNA |  | Oncogenic signature |  |
| --- | --- | --- | --- | --- | --- | --- | --- |
| 14q32 | - | Intrinsic prothrombin activation | + | MIR296 |  | MEK UP.V1 UP | - |
| 2q12 |  | Protein digestion and absorption | + | MIR34A, MIR34C, MIR449 |  | P53 DN.V1 DN | - |
| 5q35 | + | ECM-receptor interaction | + | MIR518B, MIR518C, MIR518D | + | STK33 SKM DN |  |
| 8p22 | + | Steroid biosynthesis | - | MIR135A, MIR135B | - | SINGH KRAS DEPENDENCY | - |
| 21q22 | + | Glycosphingolipid biosynthesis lacto and neolacto series | - | MIR518F, MIR518E, MIR518A | + | LEF1 UP.V1 DN | - |
| 8p21 | + | PPAR signaling pathway | - | MIR299 | + | P53 DN.v1 DN | + |
| 15q11 | + | Glycosaminoglycan biosynthesis - heparan sulfate / heparin | + | MIR339 | + | TGFB UP.V1 DN | + |
| 5q12 | + | Fatty acid degradation | - | MIR520A, MIR525 | - | ATF2 S UP.V1 UP | - |
| 9p22 | + | Fc gamma R-mediated phagocytosis | - | MIR26A, MIR26B | - | IL21 UP.V1 DN | + |
| 4q34 | - | Sulfur metabolism | - |  |  | ATF2 UP.V1 DN | + |
| 7q33 | + |  |  |  |  | MTOR UP.N4.V1 UP |  |
| 3p22 | - |  |  |  |  | EGFR UP.V1 DN | - |
| 10p15 | - |  |  |  |  | SIRNA EIF4GI UP | - |
| 4q32 | + |  |  |  |  | IL15 UP.V1 DN | + |
| 6q22 | + |  |  |  |  | ATF2 S UP.V1 DN | + |
|  |  |  |  |  |  | MYC UP.V1 DN |  |
|  |  |  |  |  |  | RAF UP.V1 DN | + |
|  |  |  |  |  |  | EIF4E DN | + |
|  |  |  |  |  |  | MEL18 DN.V1 DN | - |
|  |  |  |  |  |  | PRC2 EED UP.V1 UP | + |
|  |  |  |  |  |  | IL2 UP.V1 DN | + |
|  |  |  |  |  |  | LTE2 UP.V1 UP | - |

### Pharmacogenomics analysis to identify drugs that can potentially reverse PARP inhibitor resistance

Heterogeneity of PARPi response in the cell line data prompted us to use the molecular response signature to discover pharmaceuticals which may hold therapeutic potential in PARPi resistant tumors. We used the recently described PharmacoGx [26] analytical tool via the PharmacoDB [27] interface to analyze PARPi response across multiple large scale pharmacogenomic datasets. First, the entire set of drug interactions with the PARP response signature genes in all cell lines contained in the database was used to prioritize drugs with a highly significant (p < 0.001) interaction and reasonably large effect size (regression coefficient > |0.25|), returning a list of 35 drugs. We then compared the *in vitro* sensitivity of these 35 drugs to olaparib and rucaparib in five cancer types postulated to be at least partly sensitive to PARPi’s (breast, lung, ovarian, Ewing’s sarcoma, and osteosarcoma) by obtaining drug sensitivity measures for all available cell lines derived from the five cancer types. The cell lines were then ranked by their median IC50 value for olaparib and rucaparib, and the first and fourth quartiles of cell lines were labelled as clearly sensitive and clearly resistant. For the 35 drugs the median IC50 across resistant cell lines of each cancer type was obtained, and the 21 drugs with a median IC50 less than 10 µM for resistant cell lines of one or more cancer types are presented in Table 6. It was noteworthy that six of these drugs inhibit key enzymes in the oncogenic signatures identified in the genset enrichment analysis (Table 5, above), and four have been shown by others to synergize with PARPi’s *in vitro* (afatinib [28], crizotinib [29], erlotinib [30], and trametinib [31]) suggesting this proof of principle analysis identifies drugs which should be further investigated for the treatment of PARPi resistant tumors. Additionally, using a slightly relaxed effect size filter (regression coefficient > |0.15|) and same stringent significance cut off (p < 0.001) we found that enzastaurin, a PKCβ inhibitor previously reported to synergize with PARPi’s [32], was also identified for predictive interaction with our transcriptomic markers.

**Table 6.** Integrative Pharmacogenomic Analysis.

| Drugs | Breast |  |  | Lung |  |  | Ovarian |  |  |
| --- | --- | --- | --- | --- | --- | --- | --- | --- | --- |
|  | IC50 All | IC50 R | FC (ruc) | IC50 All | IC50 R | FC (ruc) | IC50 All | IC50 R | FC (ruc) |
| austocystin d | 0.652 | 0.913 | 47.332 | 1.228 | 1.135 | 8.029 | 1.295 | 0.156 | 2025.336 |
| ***afatinib*** | 2.123 | 2.356 | 18.339 | 4.434 | 3.397 | 2.683 | 6.154 | 5.440 | 58.253 |
| bosutinib | 4.304 | 5.643 | 7.657 | 5.485 | 4.529 | 2.013 | 4.283 | 4.861 | 65.191 |
| BRD-A86708339 | 0.506 | 0.398 | 108.506 | 1.918 | 2.620 | 3.478 | 1.755 | 1.755 | 180.581 |
| BRD-K30748066 | 1.183 | 1.183 | 36.533 | 2.575 | 4.422 | 2.061 | 0.734 | - | - |
| BX-795 | 3.950 | 7.576 | 5.703 | 3.320 | 2.417 | 3.771 | 2.398 | 2.751 | 115.207 |
| ***crizotinib*** | 7.025 | 8.072 | 5.352 | 5.735 | 7.258 | 1.256 | 8.911 | 8.256 | 38.387 |
| cytarabine | 2.281 | 6.176 | 6.996 | 1.644 | 2.749 | 3.316 | 1.686 | 2.646 | 119.785 |
| ***erlotinib*** | 32.450 | 37.318 | 1.158 | 12.381 | 8.109 | 1.124 | 14.725 | 10.461 | 30.296 |
| GSK1070916 | 10.451 | 11.733 | 3.682 | 40.708 | 6.801 | 1.340 | 66.090 | 1337.803 | 0.237 |
| ***trametinib*** | 22.735 | 60.821 | 0.710 | 0.438 | 7.240 | 1.259 | 0.165 | 2.057 | 154.056 |
| lapatinib | 4.740 | 3.154 | 13.700 | 12.283 | 9.771 | 0.933 | 11.446 | 12.411 | 25.535 |
| midostaurin | 0.533 | 0.575 | 75.088 | 0.510 | 0.679 | 13.421 | 0.430 | 0.979 | 323.712 |
| panobinostat | 0.107 | 0.096 | 449.380 | 0.070 | 0.061 | 150.348 | 0.081 | 0.112 | 2836.438 |
| *PD-153035* | 19.993 | 6.227 | 6.938 | 24.179 | 22.532 | 0.405 | 39.462 | 67.372 | 4.704 |
| *PD-0325901* | 14.347 | 20982.228 | 0.002 | 1.403 | 2.595 | 3.512 | 1.699 | 1.075 | 294.740 |
| sunitinib | 8.460 | 17.771 | 2.431 | 8.934 | 8.759 | 1.041 | 9.197 | 7.190 | 44.074 |
| TAE684 | 4.398 | 5.694 | 7.588 | 2.576 | 2.705 | 3.370 | 4.266 | 4.046 | 78.322 |
| *temsirolimus* | 4.455 | 3.483 | 12.406 | 2.309 | 0.616 | 14.796 | 0.960 | 15.901 | 19.931 |
| thapsigargin | 0.018 | 0.043 | 1013.982 | 0.018 | 0.022 | 414.276 | 0.020 | 0.023 | 13930.285 |
| tozasertib | 20.303 | 21.077 | 2.050 | 7.510 | 11.763 | 0.775 | 8.331 | 0.049 | 6491.125 |

| Drugs | Ewing's |  |  | Osteosarcoma |  |  | Pan-Cancer |  |  |
| --- | --- | --- | --- | --- | --- | --- | --- | --- | --- |
|  | IC50 All | IC50 R | FC (ola) | IC50 All | IC50 R | FC (ola) | IC50 All | IC50 R | FC (ruc) |
| austocystin d | 0.020 | - | - | 51.507 | - | - | 0.802 | 1.078 | 167.079 |
| ***afatinib*** | 13.516 | - | - | 9.495 | 2.746 | 1043.403 | 3.900 | 3.348 | 53.774 |
| bosutinib | 4.196 | - | - | 6.598 | 6.672 | 429.374 | 4.383 | 4.800 | 37.514 |
| BRD-A86708339 | 1.052 | - | - | 4.388 | 63.774 | 44.920 | 1.747 | 2.530 | 71.176 |
| BRD-K30748066 | 17.737 | - | - | 12.872 | 24.431 | 117.257 | 1.902 | 4.422 | 40.718 |
| BX-795 | 2.245 | 42.812 | 28.737 | 3.452 | 3.656 | 783.647 | 2.939 | 3.356 | 53.657 |
| ***crizotinib*** | 2.198 | 7081.577 | 0.174 | 5.507 | 5.149 | 556.401 | 6.499 | 7.770 | 23.173 |
| cytarabine | 0.297 | - | - | 0.677 | 3.079 | 930.373 | 1.477 | 2.963 | 60.777 |
| ***erlotinib*** | 8.090 | - | - | 36.789 | 12.054 | 237.655 | 19.945 | 15.173 | 11.867 |
| GSK1070916 | 0.575 | 0.608 | 2024.649 | 11.385 | - | - | 10.433 | 10.440 | 17.247 |
| ***trametinib*** | 382.041 | - | - | 0.897 | 9.469 | 302.547 | 1.814 | 22.991 | 7.832 |
| lapatinib | 10.973 | - | - | 13.833 | 112.986 | 25.355 | 10.501 | 8.944 | 20.132 |
| midostaurin | 0.470 | 0.340 | 3620.918 | 0.349 | 0.332 | 8621.515 | 0.498 | 0.604 | 298.176 |
| panobinostat | 0.023 | - | - | 0.112 | 0.192 | 14884.391 | 0.080 | 0.080 | 2260.037 |
| *PD-153035* | 22.649 | - | - | 39.954 | 44.258 | 64.727 | 25.223 | 24.497 | 7.350 |
| *PD-0325901* | 0.026 | - | - | 5.199 | 395.942 | 7.235 | 2.510 | 6.774 | 26.579 |
| sunitinib | 3.341 | - | - | 6.578 | 4.113 | 696.511 | 8.705 | 8.534 | 21.100 |
| TAE684 | 0.663 | 27.199 | 45.234 | 1.143 | 1.066 | 2687.037 | 2.844 | 3.394 | 53.047 |
| *temsirolimus* | 0.009 | - | - | 0.506 | 0.242 | 11853.130 | 2.967 | 1.125 | 160.005 |
| thapsigargin | 0.007 | 0.003 | 439669.837 | 0.003 | 0.023 | 124818.344 | 0.016 | 0.023 | 7914.719 |
| tozasertib | 2.225 | 38.784 | 31.722 | 4.504 | 29.366 | 97.551 | 11.974 | 16.029 | 11.233 |

## Discussion

PARP inhibitors have significantly improved progression-free and overall survival for patients with BRCA positive ovarian [10] and breast cancer [11], [12]. The clinical benefit, however, is not restricted to patients with BRCA mutation mediated HR deficiency and in clinical trials, the survival advantage is also observed in patients with wild type BRCA genes [33], [34]. Moreover, *in vitro* studies indicate that PARP inhibitors have broad activity, extending beyond clinically approved indications in BRCA mutated ovarian and breast cancer patients [16]. In this study, we therefore investigated if a gene expression signature could be used to predict the clinical response of PARP inhibitors in both cancer cell lines and patients.

Based on the expression profile of a subset of marker genes from cell lines treated with olaparib, we were able to predict with high accuracy response of an independent PARP inhibitor, rucaparib, as well resistance and sensitivity of cell lines from a completely independent study. Furthermore, the model outperformed our previously published “BRCAness” signature when tested on multiple different cell lines, indicating that the signature identified in this study is not restricted to the pathways captured by the BRCAness signature.

The signature also predicted with high accuracies the clinical response of patients treated with platinum-based chemotherapy, which was used as a surrogate for PARP inhibitor response, in patients with ovarian cancer and osteosarcoma. This indicates that the gene expression profile covers not only the broad biology underlying the PARP inhibitor response but also suggests that such an approach could be used on patient-derived samples.

There are, at the moment, no available clinical data that would enable us to confirm how well the model would perform when actual patients are on PARP inhibitors. However, since the clinical decision to start PARP inhibitor in ovarian cancer is made when the patient is either on or has just completed platinum-based chemotherapy, this might more closely approximate current clinical practice than a pure predictor for PARP inhibitor response. That said, the predictive performance of such a signature should ultimately be tested in patients only treated with a PARP inhibitor as well.

Given that the clinical utility of PARP is now being tested across cancers without known HRD we suggest that such a predictive model would be essential to identify the subgroup of patients who might benefit from the PARP inhibitors, especially in cancers where the clinical benefit is most likely to be restricted a subset of patients. Our results indicate that cancer subsets that were previously thought not to be susceptible to PARP inhibitors may still benefit from treatment with these drugs with careful selection of patients based on tumor expression profile. The data from our study show that the response predictive gene list is highly reproducible in various independent cancer data sets, including subsets that were previously thought to be unsusceptible to PARP inhibitors.

Our results also demonstrate the value of principled and sophisticated analysis of large genomic and drug response datasets such as the CCLE [35], [36] and the GDSC [18], [37], [38]. While some concerns were previously raised about some aspects of technical reproducibility of these drug response screening efforts [39], [40], our study demonstrates that despite those potential issues, these datasets, when analyzed carefully and with adequate steps to minimize artifacts and noise, can be very valuable in the effort to reveal new therapeutic applications and develop response predictors. In this context, another study recently utilized expression and drug response data successfully develop and test a PARP inhibitor predictor [32]. That report utilized a different set of gene expression data from our study (CCLE vs GDSC) and employed different analytical algorithms and somewhat different definitions of response vs non-response. Likely due to all these differences in study approach, only a few of the top marker genes overlapped with the multi gene signature we describe here.

However, both studies converge on similar overall conclusions on the power of gene profiles to predict response to PARP inhibitors and aid in developing novel therapies for PARP inhibitor resistance. As has been the case for other phenotypes (such as breast cancer prognostic microarray signatures), it is possible that the different marker panels reflect different aspects of the same biological phenotype which is characterized of a large number of differentially expressed genes, as shown in our analysis too.

Our pharmacogenomic bioinformatic analysis of these large transcriptional differences between cell lines sensitive and resistant to PARPi’s revealed drugs which may have a rationale for synergy with PARPi’s. Furthere, this analysis identified four drugs for which pre-clinical evidence of synergistic interactions with PARPi’s already exists, namely afatinib [28], crizotinib [29], erlotinib [30], and trametinib [31]. This suggests the drugs from our analysis are candidates for further study, particularly in the subset of patients predicted to be unresponsive to PARPi therapy by the PARP response signature. Clinical trials often reveal that only a subset of patients respond to treatment with novel molecularly driven therapies, and recent PARPi studies in Ewing sarcoma underscore this point [41]. In this context, expression read outs, such as the signature presented in this study, can potentially become useful clinical markers to prioritize these new treatments to a subset of patients more likely to respond, and may be used to also identify drugs with possible synergy for further testing in the clinical setting.

Techniques used to detect homologous recombination deficits, such as RAD51 foci formation assays and genomic scar scoring, have been proposed as possible tools to predict response to PARPi’s [42]. Despite their proposed utility, RAD51 foci detection can only be performed in tumors after exposure to DNA damaging agents, or *ex vivo* DNA damage, and genomic scar biomarkers have so far been limited by a low positive predictive value [43]. Further work should be conducted to determine if these methods can complement a transcription-based assay of PARPi response, which has the additional advantage that it can be performed prior to the decision to treat with the drugs.

Multi-gene predictors, such as the one developed in this study, need further refinement and validation, and future prospective studies are required to confirm the clinical utility of a predictive model. With the wide availability of RNA sequencing, we think a predictive model based on RNA expression, in conjunction with DNA sequencing approaches, would be feasible for future clinical trials and would help extend the utility of PARP inhibitors to an expanding array of patient subgroups in many different types of cancers.

## Methods

### *In vitro* PARP inhibitor response

Cell line drug response and transcription data used to generate the PARPi response signature was obtained from the GDSC [37] dataset https://www.cancerrxgene.org/downloads. The median IC50 of olaparib and rucaparib across all cell lines tested in the dataset was used to classify each cell line as sensitive or resistant to olaparib and rucaparib, separately. To minimize technical artifacts and prioritize the drug class as opposed to specific compound effect, cell lines were required to be classified concordantly for olaparib and rucaparib response in order to be included in the analysis. An independent gene transcription dataset of breast cancer cell lines treated with olaparib was obtained from a previously published study [21], which was used as entirely independent in vitro dataset to validate the PARPi response signature.

### Human ovarian cancer and osteosarcoma gene expression datasets

Clinical and gene expression data for 456 ovarian cancer cases treated with cisplatin were obtained from The Cancer Genome Atlas data portal (https://portal.gdc.cancer.gov/) [22]. We previously generated the clinically annotated osteosarcoma expression profiling dataset [23], which is accessible through GEO Series accession number GSE39055.

### Unsupervised hierarchal clustering and standard univariate tests

Differential analysis (class comparison) between two groups of samples was performed by a permutation-based t-test with standard False Discovery Correction for multiple testing [44]. Positional genesets, miRNA target genesets, and oncogenic signature genesets were downloaded from the Molecular Signatures Database (http://software.broadinstitute.org/gsea/msigdb). BioCarta genesets were downloaded via the Cancer Genome Anatomy Project (http://cgap.nci.nih.gov/Pathways) and KEGG pathways via the KEGG.db R package (http://www.bioconductor.org/packages/release/data/annotation/html/KEGG.db.html). Unsupervised hierarchal clustering [20] was performed with the centered correlation and average linkage method. Associations between two categorical variables were evaluated with two-tailed chi-square/Fisher’s exact test. Cluster-based group associations with survival were assessed by Kaplan-Meier analysis and the log rank test for significance. Pearson’s r statistic was used to evaluate continuous variable correlations.

### Response/Survival prediction and ROC analysis

Binary prediction accuracy of the PARP inhibitor signature (sensitive / resistant) was predicted using a 3-fold or leave one out cross validated logistic lasso regression [45] and continuous response prediction was performed with least angle regression (LARS) [46] as previously described. The supervised principal components survival prediction method was used to generate models for patient survival and perform leave-one-out cross validated performance assessment [47]. Receiver operating characteristics (ROC) and area under the curve (AUC) analysis was performed per standard methodology using a continuous response probability index generated by a Bayesian compound covariate prediction algorithm [48].

### Geneset enrichment analysis

Geneset enrichment analysis [49] for the association of global expression profiles (filtered for the lowest 20% variance genes) with PARP inhibitor sensitivity was performed with the functional class scoring method [50] applying the LS/KS test with a permutation p value less than 0.05 by both tests used to identify genesets with enriched differential expression. Positional genesets, miRNA target genesets, and oncogenic signature genesets were downloaded from the Molecular Signatures Database (http://software.broadinstitute.org/gsea/msigdb).

BioCarta pathway genesets were downloaded from the Cancer Genome Anatomy Project (http://cgap.nci.nih.gov/Pathways) and KEGG pathways were obtained using the KEGG.db R package (http://www.bioconductor.org/packages/release/data/annotation/html/KEGG.db.html).

### Integrative pharmacogenomic analysis

The 50 gene PARP inhibitor response signature was analyzed for drug interaction discovery using the PharmacoGx [26] R package through the PharmacoDB [27] interface. Specifically, 43 unique genes comprising the signature were individually tested for association with drug response across seven large datasets using stringent selection criteria for effect size (regression coefficient > |0.25|) and significance (two-sided t-test p < 0.001) resulting in 35 drugs with at least one significant interaction with a gene from the PARPi response signature. Whenever we tested a drug hypothesis derived by previously published data, external to this analysis, we used a slightly less restrictive, but still stringent selection criteria for effect size (regression coefficient > |0.15|) with the same significance cut off (two-sided t-test p < 0.001).

*In vitro* drug sensitivity metrics were then obtained for the 35 drugs with cell lines corresponding to the five tissue types of interest; namely breast (87 cell lines), lung (221 cell lines), ovarian (64 cell lines), osteosarcoma (15 cell lines), and Ewing’s sarcoma (25 cell lines). We elected to use the median IC50 for groups of cell lines in order to avoid outlier effects. First, the median IC50 dose response metric was calculated for each drug across the cell lines to obtain a “pan-cancer” IC50 value for each drug. Median IC50 values for each of the five cancer types were then calculated separately to observe tissue-type specific drug interactions. To identify cell lines resistant and sensitive to the PARP inhibitors olaparib and rucaparib in the PharmacoDB database, median IC50’s for olaparib and rucaparib were obtained for the subset of cell lines for which drug response data was available. For each cell line, the median response to both olaparib and rucaparib was calculated. Cell lines were ranked by the median IC50 response, and the 1^st^ and 4^th^ quartiles of the list were used to define sensitive and resistant cell lines, respectively. Within the separate sensitive and resistant cell line groups, median IC50 values for each drug were calculated. As a final filtering step, only drugs with a median IC50 less than 10 µM in at least one tissue type’s subset of PARPi resistant cell lines were included in our final list of 21 drugs.

### Statistical Software

The NCI BRB-ArrayTools v4.6.0 [51], R (version 3.4.3), and SPSS v 24 software were used.

## Data Availability

The data analyzed in this study are publicly available at the Genomics of Drug Sensitivity in Cancer resource page (https://www.cancerrxgene.org/downloads/bulk_download), the TCGA GDC data portal (https://portal.gdc.cancer.gov/projects/TCGA-OV), ArrayExpress (E-TABM-157), and Gene Expression Omnibus (GSE:39058).

https://portal.gdc.cancer.gov/projects/TCGA-OV

https://www.ncbi.nlm.nih.gov/bioproject/PRJNA169852

## Supporting Information

**Supplementary Table 1**. Cell lines included in the analysis.

**Supplementary Table 2**. Genes differentially expressed (FDR < 0.05) between olaparib sensitive and resistant cell lines.

**Supplementary Table 3**. Osteosarcoma cell line response to the PARPi’s olaparib and rucaparib.

**Supplementary Table 4**. Geneset analysis results with LS and KS permutation p values for enriched genesets in the olaparib and rucaparib datasets.

